# Epidemiology of Epstein-Barr Virus infection and Infectious Mononucleosis in the United Kingdom

**DOI:** 10.1101/2020.01.21.20018317

**Authors:** Ashvin Kuri, Benjamin Meir Jacobs, Nicola Vickaryous, Julia Pakpoor, Jaap Middeldorp, Gavin Giovannoni, Ruth Dobson

## Abstract

**Background:** Epstein-Barr Virus (EBV) is a ubiquitous gamma-herpesvirus with which ∼95% of the healthy population is infected. EBV infection has been implicated in a range of haematological malignancies and autoimmune diseases. Delayed primary EBV infection increases the risk of subsequent complications. Over recent decades, the age of primary EBV infection has become later, largely due to improved sanitation and living conditions.

**Methods and findings:** First, we conducted a sero-epidemiological survey of healthy volunteers between 0 and 25 years old to assess prevalence of detectable anti-EBV antibodies. 1982 of 2325 individuals (85.3%) were EBV seropositive. EBV seropositivity increased monotonically with age, and increased more among females than males during adolescence (ages 10 – 15). Second, we conducted a retrospective review of Hospital Episode Statistics to determine changes in Infectious Mononucleosis (IM) incidence over time. Between 2002 and 2013, the incidence of IM (derived from hospital admissions data) increased. We then conducted a large case-control study of 6306 prevalent IM cases and 1,009,971 unmatched controls extracted from an East London GP database to determine exposures associated with IM. Exposures associated with lower risk of IM were elevated BMI (Overweight OR 0.80 [0.75 to 0.86], obese OR 0.63 [0.57 to 0.70]), non-white ethnicity (Black OR 0.21 [0.18 to 0.23], Asian OR 0.14 [0.13 to 0.16], Other ethnicity OR 0.22 [0.19 to 0.25]), and a history of smoking (OR 0.87 [0.83 to 0.92]), whereas affluence was associated with a higher risk of IM (per increase in IMD decile OR 1.15 [1.13 to 1.17]. Finally, we used ELISA to determine antibody responses to common pathogens and vaccine antigens among EBV-seronegative individuals. EBV-seronegative donors did not display diminished serum antibody responses to pertussis, rubella, or varicella compared to EBV-seropositive donors.

**Conclusions:** In this study we make several important observations on the epidemiology of EBV infection in the UK. We find that overall EBV seroprevalence in the UK appears to have increased, and that the sharp increase in EBV seropositivity takes places earlier among females than males. We find that the incidence of IM requiring hospitalisation is increasing. We find that exposures associated with prevalent IM in a diverse population include white ethnicity, affluence, lower BMI, and never-smoking, and these exposures interact with each other. Lastly, we provide pilot evidence suggesting that antibody responses to vaccine and encountered pathogens do not seem to be diminished among EBV-seronegative individuals, which is a theoretical counter-argument to developing EBV vaccines. Our findings could help to inform vaccine study designs in efforts to prevent IM and late complications of EBV infection, such as Multiple Sclerosis.

**Key messages:**

- Epstein-Barr Virus (EBV) is a ubiquitous virus which infects over 95% of the world’s population. The majority of infection is silent and without consequence. In a subset of individuals, EBV is thought to play a role in the pathogenesis of autoimmune disease and haematological cancers.
- During childhood and adolescence, EBV seroprevalence increased monotonically with age from 0-5 (67.8% females, 72.0% males) to 20-25 (96.4% females, 95.5% males)
- The incidence of Infectious Mononucleosis (IM) leading to hospital admission has increased over the past decade
- Exposure associated with IM in a large, diverse East London cohort (n>1,000,000) were low BMI, never-smoking, white ethnicity, and affluence.

## Introduction

Epstein-Barr Virus (EBV) is a ubiquitous gamma-herpesvirus with which ∼95% of healthy adults are infected^1^. EBV is the commonest causative agent of Infectious Mononucleosis (IM), a triad of pharyngitis, lymphadenopathy and fever in the context of acute EBV infection^2^. EBV infection is also implicated in a range of haematological malignancies (Burkitt’s lymphoma, gastric lymphoma, and Non-Hodgkin lymphoma), and has been strongly associated with autoimmune diseases such as Multiple Sclerosis (MS).

EBV seroprevalence increases monotonically with age, and tends to be higher among females, non-Caucasian ethnic groups, and people living in socio-economically deprived households at any given age^3,4^. Delayed primary infection with EBV is a risk factor for IM, with later age at EBV infection conferring both a higher risk of IM and a higher risk of severe features^2^. Concerningly, overall EBV seroprevalence rates among children and adolescents appear to be decreasing in some populations, implying that the population at risk of complicated primary EBV infection is increasing ^4–6^.

Various strands of evidence suggest a pathogenic role for EBV infection in MS: EBV seronegativity is exceptionally rare in people with MS, EBV seropositivity is higher in people with MS, anti-EBV antibody titres predict the risk of MS, and prior IM increases the risk of subsequent MS^7–10^. Understanding the epidemiology of EBV infection during childhood and adolescence is essential for maximising the efficacy and safety of vaccine studies^11^ and in order to inform power calculations for future potential interventional studies.

Previous EBV vaccine studies have demonstrated efficacy against symptomatic IM but not EBV seroconversion^12^; however newer generation vaccines are being developed to prevent seroconversion^13^. Prior to vaccine development, there is an urgent need to better understand the current epidemiology of EBV transmission, and potential associations with both early and late seroconversion.

We therefore set out to update and establish current EBV seroprevalence rates across the UK, establish the potential effect of changing EBV seroprevalence on rates of Infectious Mononucleosis (IM), and examine predictors of late EBV infection (IM) in a large and highly diverse primary care cohort. Finally, we used a small cohort of truly EBV-negative samples from adults to explore whether EBV seronegativity is associated with a reduced immune response to other vaccine-preventable and/or ubiquitous infections.

## Methods

### Samples

Serum samples used to investigate the prevalence of EBV-seropositivity across the UK population were requested from Public Health England Seroepidemiology Unit (SEU). The SEU houses a serum repository formed from residual samples from diagnostic testing across NHS laboratories in England. Samples are anonymised but retain data pertaining to age, sex, date and laboratory of collection. A request was submitted for 2500 samples spread across age groups and English geographical area (London, South West, West Midlands, North West, North East) dating from 2016. A total of 2366 blood serum samples were available for analysis from the SEU.

A second cohort of 21 adult EBV-seronegative and 42 age and gender matched EBV-seropositive samples, were collected from a Netherlands cohort to compare the immunological properties associated with serostatus. EBV seronegativity was defined using an in-house VCA-IgG and EBNA1-IgG ELISA, followed by confirmation with an IgG immunoblot against VCA, EA, and EBNA^14^.

All serum samples were stored at −80C until analysis, and freeze thaw cycles kept to a minimum. No samples had >3 freeze thaw cycles prior to analysis.

### Enzyme-linked immunosorbent assay (ELISA)

The anti-Epstein Barr virus (EBV-VCA) IgG Human ELISA kit (Abcam, cat no. ab108730) was used for qualitative determination of IgG antibodies against EBV viral capsid antigen (VCA) in human serum. IgG VCA antibodies peak two to four weeks post EBV infection, and persist for life, serving as a marker for recent or historic EBV infection. Serum samples were diluted 1:100 immediately prior to use and assayed according to the manufacturer’s protocol. Positive, negative and cut-off controls were run in duplicate; samples were run in singlicate. Any sample falling within the +/-10% cut-off range were repeated in duplicate; repeat samples remaining in the +/-10% cut-off range were not included in the final analysis.

Quantitative measurement of IgG directed against *Rubella* and *Varicella zoster virus* (VZV) and semi-quantitative measurement of IgG against *Bordetella pertussis* was carried out on matched EBV positive and negative samples using ELISA kits (cat no. KA0223, KA1456, and KA2090 respectively; Abnova Corporation, Taipei City, Taiwan). Briefly, serum samples were diluted 1:100 and run in duplicate according to manufacturer’s instructions. Assay validity was confirmed using control samples provided by the manufacturer in all cases.

### Hospital Episode Statistics (HES)

Hospital admissions with infectious mononucleosis (IM) recorded as either a primary or secondary diagnostic code during admission were examined using Hospital Episode Statistics (HES). HES incorporate every episode of hospital day-case or overnight inpatient care in National Health Service (NHS) hospitals^15^. The Oxford record-linkage group undertook record-linkage to construct absolute numbers of recorded hospital attendances with IM for each year 2002-2013. Infectious mononucleosis was defined as ICD-10 code B27.9 and ICD-9 code 075. Absolute numbers of cases of IM per age band were converted to rates per 100,000 person-years for each 5 year age band for analysis. As this was a descriptive study no reference or control cohort was required.

### Primary care data analysis

The East London Primary Care dataset consists of de-identified primary care (General Practitioner, GP) healthcare records from all GP practices using the EMIS electronic healthcare records system (https://www.emishealth.com/) across four clinical commissioning groups (CCGs) in east London - Hackney, Newham, Tower Hamlets and Waltham Forest. To determine environmental and ethnic exposures associated with IM risk, we conducted a large case-control study using East London GP database records. We extracted available demographic details for all participants within the database.

Infectious mononucleosis was defined using codes for ‘Glandular fever’, ‘confirmed glandular fever’, ‘gammaherpesviral mononucleosis’, ‘infective mononucleosis’, ‘infectious mononucleosis’, and ‘Pfeiffer’s disease’. Self-declared ethnicity at GP registration was used in these analyses.

Ethnicity codes used within EMIS map directly onto four main Office for National Statistics (ONS) ethnicity codes used in the UK census: White, South Asian, Black, and other. Smoking status was taken at the latest date recorded. An exception to this was where an earlier status was recorded as current/ex-smoker and later status never smoker, re-coded as ex-smoker. Never smoker was the reference group. Earliest recorded height/weight was used. Where height was outside the range 85-250cm, weight outside the range 9-250kg, or BMI outside the range 10-50kg/m^2^, these data were classified as missing/unknown data, as such values were felt to be implausible. Where height/weight were unavailable but there was a record of obesity as a diagnosis, BMI was categorised as obese.

IMD is a geographical measure, based on socio-economic terms, including income, employment, education, health, crime, housing and environment. IMD raw scores for each lower layer super output area (LSOA) were assigned to deciles derived from the national distribution using ONS data and treated as a continuous variable. Decile 1 represents the most deprived 10% of the population, and decile 10 represents the least deprived 10% (most affluent).

### Ethical approval

SEU sample analysis was covered by SEU ethical approvals which include consent for use in research (reference 05/Q0505/45); analysis of the EBV seronegative cohort was covered by internal ethical approval from VU University, Amsterdam. Primary and secondary care data analysis was performed on fully anonymised unlinked records and separate ethical approvals and additional consent were not required.

### Statistical analysis

To examine the determinants of EBV serostatus and IM we used univariate and multivariate logistic regression. The strength of association between a variable and EBV serostatus was determined using the likelihood ratio of the full model vs that containing only confounders. Differential rates of seropositivity to other infections was compared between EBV-positive and EBV-negative cohorts using Fisher’s exact test. Unless otherwise stated, results are presented as estimate followed by 95% confidence interval (CI). All analyses were conducted using R version 3.6.1 in RStudio.

## Results

### UK EBV seroprevalence

Sample demographics are displayed in supplementary table 1. 41/2366 individuals had indeterminate EBV serology results. Of the 2325 participants with definite serostatus results, there was a roughly equal gender split (51.4% female). Overall 1982/2325 (85.3%) were EBV seropositive.

EBV seroprevalence increased monotonically with age from 0-5 (67.8% females, 72.0% males) to 20-25 (96.4% females, 95.5% males, table 1, figure 1). Age band was strongly associated with EBV seropositivity (p<0.0001) in both univariate models and models adjusting for sex and region. Repeating the analysis with age as a continuous predictor yielded similar results, with each year increase in age corresponding to a 12% increase in the odds of seropositivity (OR 1.12, 95%CI 1.10-1.14, p<0.0001.

**Table 1:**
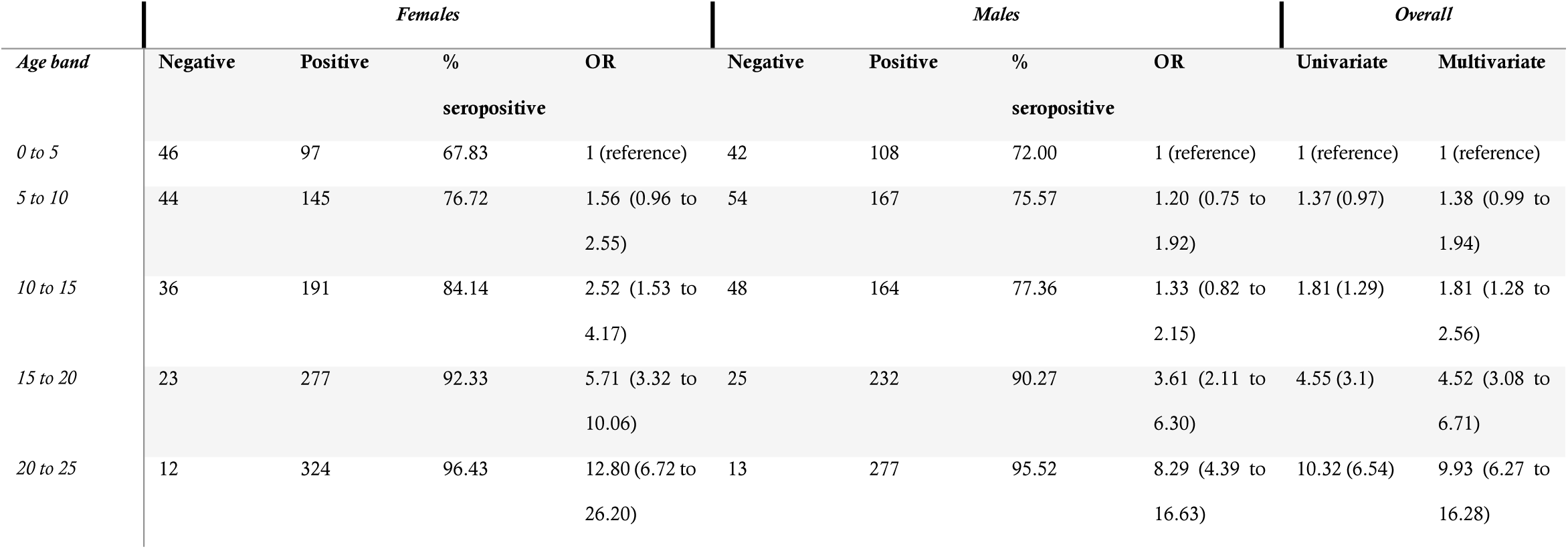
raw numbers of seropositive and seronegative individuals within each age and sex category, univariate and multivariate odds ratios for seropositivity for males, females, and combined estimates.

**Figure 1:**
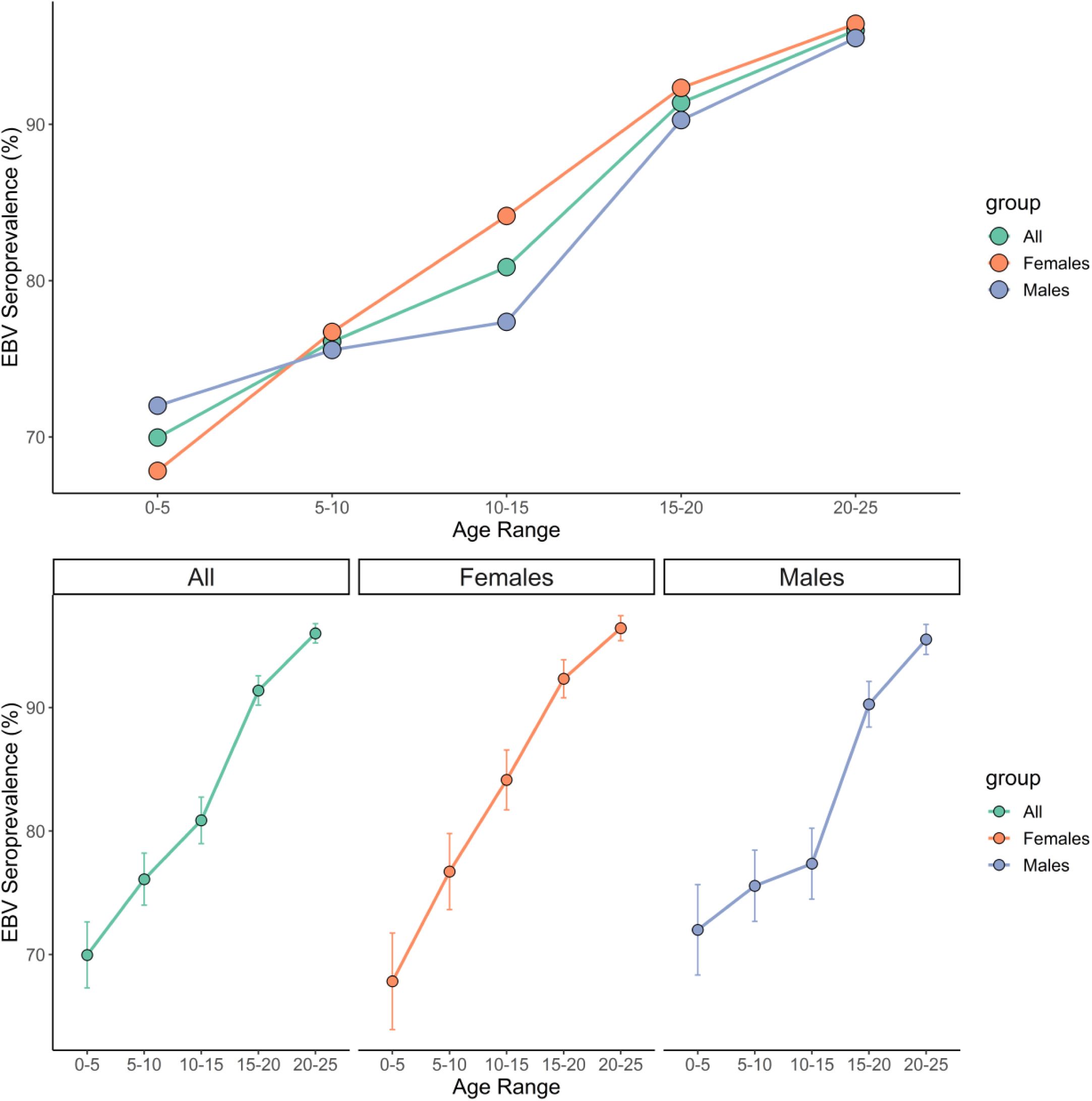
top panel – EBV seroprevalence among healthy volunteers split by sex and age range. Bottom panel – same data as top panel, presented separately with 95% confidence intervals for each sex.

Sex was not sigificantly associated with EBV seropositivity in a univariate model (OR_Male_ 0.81, 95%CI 0.64-1.02, p=0.074). This effect further dissipated on correcting for region in a multivariate model (OR_Male_ OR 0.84, 95% CI 0.67 to 1.06, p = 0.15). Region was associated with EBV seroprevalence in a univariate model, with evidence of lower seropositivity in non-London regions (p < 0.01), however this effect dissipated on correction for age and sex (supplementary table 2).

Seroprevalence estimates were similar between males and females at most age groups: a striking exception is the 10-15 age band, in which the point estimate for EBV seroprevalence was far higher among females (84.1% [79.4% to 88.9%]) than males (77.4% [71% to 83.0%), although these estimates were imprecise.

### Temporal trends in the incidence of Infectious Mononucleosis 2002 - 2013

To determine the temporal trends in the incidence of Infectious Mononucleosis, we examined Hospital Episode statistics over an 11 year period from 2002 to 2013. Over the time period studied, IM incidence as measured by hospital admission data steadily rose for males and females (supplementary table 3). There was no clear shift in sex ratio or the age distribution of incident IM cases, with 15-19 year olds accounting for the majority of diagnoses in both time periods studies (figure 2).

**Figure 2:**
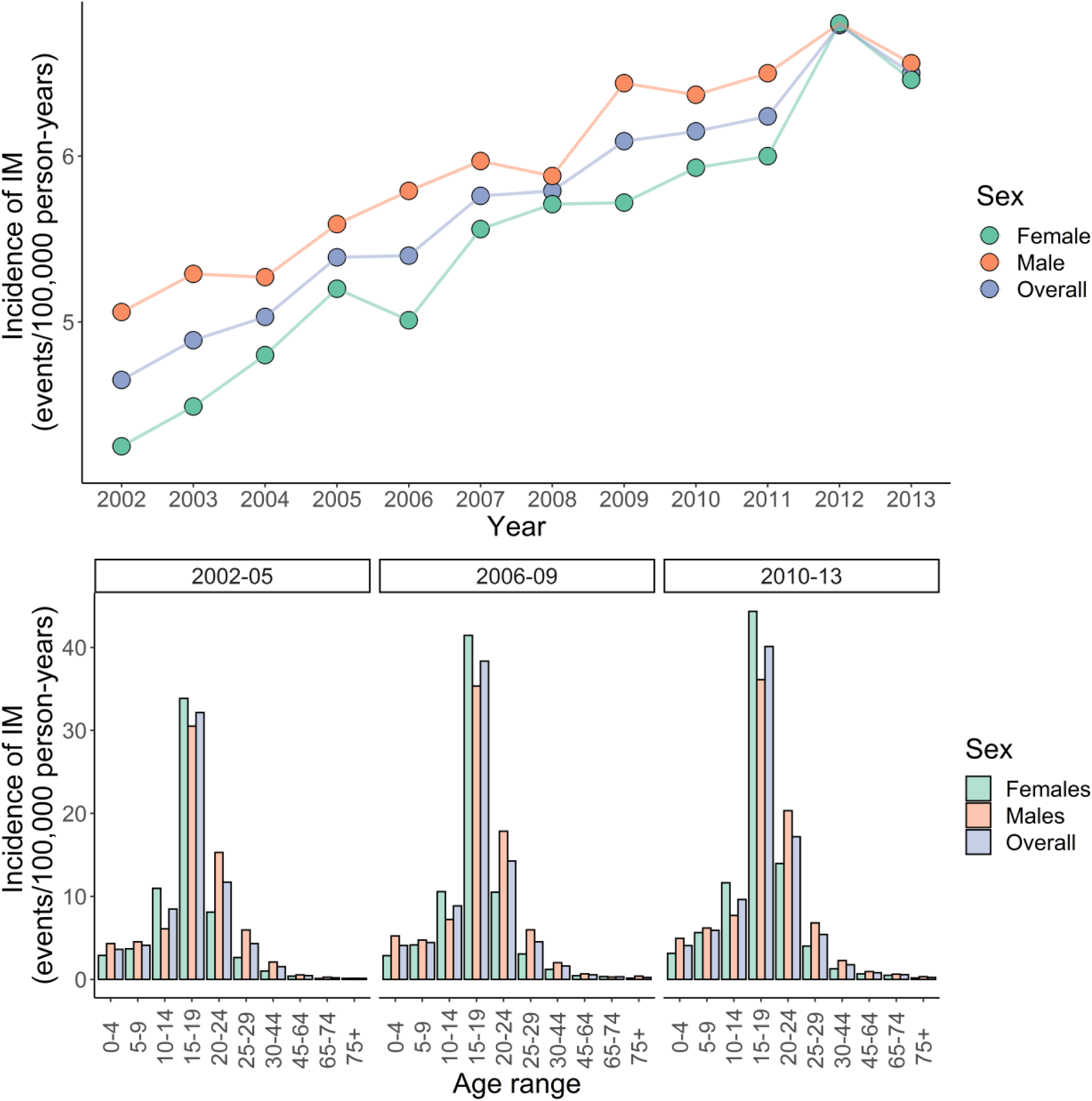
top panel – incidence of infectious mononucleosis (IM) derived from Hospital Episode Statistics between 2002 and 2013 divided by sex. Bottom panel – same data as top panel presented grouped by epochs of 3 years, presented separately for each age range.

### Exposures associated with Infectious Mononucleosis

In total, we extracted data for 6306 IM cases and 1,009,971 unmatched controls. Demographics of included participants are shown in table 2. We fitted multivariate logistic regression models using prevalent IM status (i.e. any historical episode of IM) as the outcome, and incorporating age and sex as confounders. Adding BMI, ethnicity, smoking, and IMD decile to the model all improved the model fit, suggesting that these exposures explain at least some of the variation in IM prevalence.

**Table 2:** demographics of IM cases and unmatched controls in the East London GP database, including prevalence of IM among each group.

|  | *Controls (1009971)* | *Cases (6306)* | *Prevalence (% cases)* |
| --- | --- | --- | --- |
| ***Age*** | 40.49 (15.44) | 39.19 (13.08) |  |
| ***Sex*** |  |  |  |
| *Female* | 493384 | 3193 | 0.64 |
| *Male* | 516573 | 3113 | 0.60 |
| ***Ethnicity*** |  |  |  |
| *White* | 440066 | 4865 | 1.09 |
| *Black* | 135689 | 282 | 0.21 |
| *Other* | 114712 | 283 | 0.25 |
| *S.Asian* | 217426 | 377 | 0.17 |
| ***BMI*** |  |  |  |
| *Normal* | 388300 | 3031 | 0.77 |
| *Obese* | 130458 | 543 | 0.41 |
| *Overweight* | 253323 | 1361 | 0.53 |
| *Underweight* | 136876 | 966 | 0.70 |
| ***Smoking*** |  |  |  |
| *Never smoker* | 561648 | 3396 | 0.60 |
| *Ever smoker* | 399975 | 2800 | 0.70 |
| ***IMD decile*** |  |  |  |
| *1* | 137431 | 561 | 0.41 |
| *2* | 317377 | 1692 | 0.53 |
| *3* | 303976 | 1746 | 0.57 |
| *4* | 120676 | 924 | 0.76 |
| *5* | 55535 | 628 | 1.12 |
| *6* | 20756 | 244 | 1.16 |
| *7* | 13018 | 129 | 0.98 |
| *8* | 6154 | 75 | 1.20 |
| *9* | 6532 | 89 | 1.34 |
| *10* | 335 | 8 | 2.33 |

We found evidence that these exposures are independently associated with IM, as their effects persisted in a combined model incorporating all factors as covariates. Factors inversely associated with IM (‘protective’) were elevated BMI (Overweight OR 0.80 [0.75 to 0.86], obese OR 0.63 [0.57 to 0.70]), non-white ethnicity (Black OR 0.21 [0.18 to 0.23], Asian OR 0.14 [0.13 to 0.16], Other ethnicity OR 0.22 [0.19 to 0.25]), and a history of smoking (OR 0.87 [0.83 to 0.92]). Affluence was associated with a higher risk of IM (per increase in IMD decile OR 1.15 [1.13 to 1.17], table 3).

**Table 3:** multivariate odds ratios for IM with 95% CIs (adjusted for age and sex) derived from East London GP database. P values represent empirical p values for each level of the predictor (3^rd^ column) and likelihood ratio p values (4^th^ column) represent the additional improvement in model fit on adding the predictor to a null model consisting of only age and sex.

| *Exposure* | *OR (95% CI)* | *P value* | *Likelihood ratio P* |
| --- | --- | --- | --- |
| ***Age*** | 0.99 (0.99 to 0.99) | <0.0001 |  |
| ***Sex*** |  |  |  |
| *Female* | 1 (reference) |  |  |
| *Male* | 1.01 (0.95 to 1.06) | 0.851 |  |
| ***Ethnicity*** |  |  | <0.0001 |
| *White* | **1 (reference)** |  |  |
| *Black* | 0.21 (0.18 to 0.23) | <0.0001 |  |
| *Other* | 0.22 (0.19 to 0.25) | <0.00013 |  |
| *South asian* | 0.14 (0.13 to 0.16) | <0.00012 |  |
| ***IMD Decile (2015)*** | 1.15 (1.13 to 1.17) | <0.0001 | <0.0001 |
| ***Weight*** |  |  | <0.0001 |
| *Normal weight* | 1 (reference) |  |  |
| *Obese* | 0.63 (0.57 to 0.7) | <0.0001 |  |
| *Overweight* | 0.8 (0.75 to 0.86) | <0.0001 |  |
| *Underweight* | 1.03 (0.95 to 1.11) | 0.465 |  |
| ***Smoking status*** |  |  | <0.0001 |
| *Never smoker* | 1 (reference) |  |  |
| *Ever smoker* | 0.87 (0.83 to 0.92) | <0.0001 |  |

We then looked for pairwise interactions between all significant predictors of IM status, controlling for age and gender. We considered an interaction significant if including the interaction term improved model fit at an adjusted p threshold of 0.05/6=0.00833. We observed significant pairwise interactions between ethnicity and IMD, ethnicity and weight, ethnicity and smoking, and weight and IMD (supplementary table 4). Repeating this analysis using weight as a binary measures (grouping “normal” and “underweight” individuals, and grouping “overweight” and “obese” individuals) gave similar results (data not shown). Strikingly, we noted opposite effects of smoking and body weight depending on ethnic group: the ‘protective’ effect of being overweight was reversed in Asian people, and the ‘protective’ effect of smoking was only observed in White people (figure 4).

**Figure 3:**
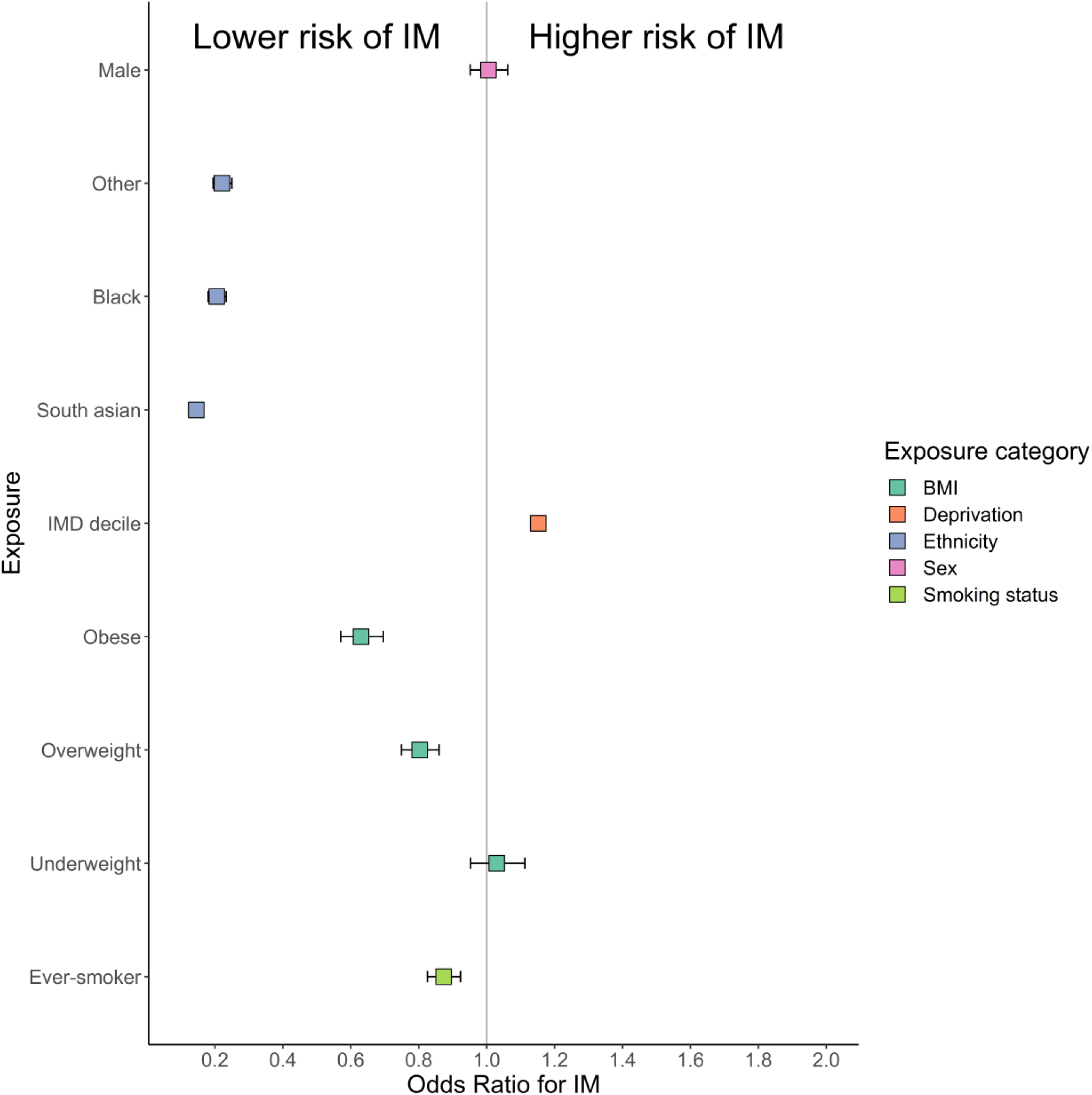
forest plot depicting exposure associations with IM in the East London GP database. Data points represent odds ratios +/- 95% confidence intervals.

**Figure 4:**
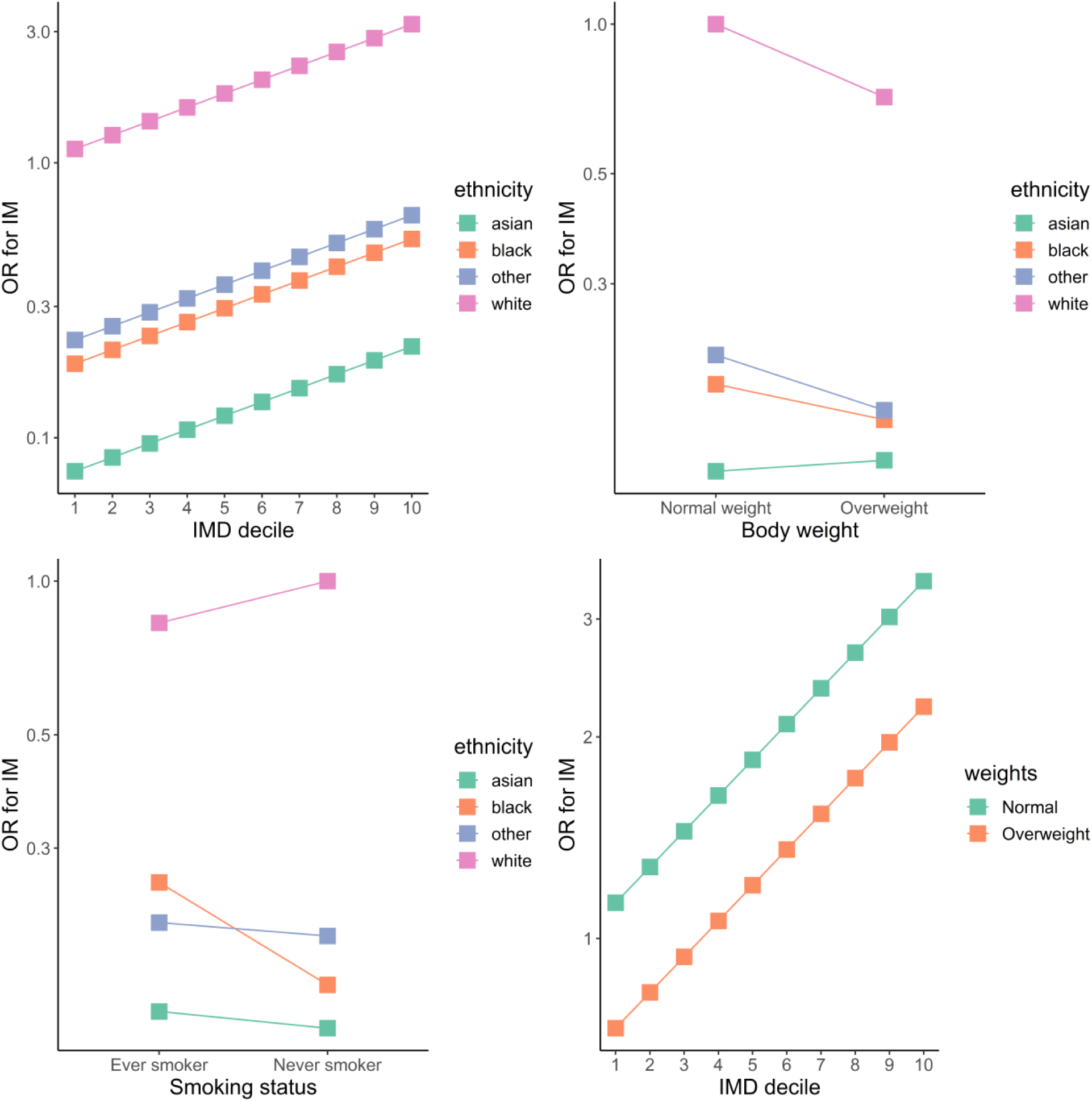
odds ratios for infectious mononucleosis derived from multivariate models incorporating interaction terms between individually significant predictors of IM status.

### EBV seronegativity is not associated with diminished serologic response to vaccine antigens

To test the hypothesis that EBV-seropositivity is biologically necessary to prime and maintain B cell memory responses to vaccine antigens, we assayed IgG against recall antigens. Sera were confirmed to be EBV-seronegative using the methods described above; matched control sera were EBV seropositive. We found no evidence of differences in Rubella, Pertussis, or Varicella serostatus between EBV seronegative and EBV seropositive individuals (data not shown).

## Discussion

In this study we demonstrate the temporal trends in EBV seroprevalence during childhood and adolescence among healthy UK volunteers, and explore the socio-demographic associations with IM (a marker of late infection). We show that the seroprevalence of EBV increases monotonically with age from a nadir of 69.9% in the 0-5 age bracket to 96.0% among 20-25 year olds. Furthermore, we show that the temporal trends in EBV seroprevalence differ between genders, with male sex conferring a slightly lower risk of EBV seropositivity during adolescence (10 - 15 years), which corresponds to a gender imbalance among hospital-coded infectious mononucleosis cases.

We show that, among a diverse East London population, exposures associated with increased IM risk include white ethnicity, normal/low body weight (being overweight is protective), lower deprivation level, and never smoking. We show evidence of interaction between smoking, body weight, and ethnicity in determining IM risk. Finally, we provide some pilot evidence suggesting that EBV seronegativity does not impair antibody responses to recall antigens.

Our results are consistent with previous studies on the determinants of EBV seropositivity. It is well-established that EBV seroprevalence increases with age^3–6,16–20^. Data from the US National Health and Nutrition Examination Surveys (NHANES) demonstrated that predictors of EBV seronegativity were male sex, white ethnicity, younger age, higher household income, lower household crowding, higher health insurance ^3,4^. A UK-based study demonstrated a dramatic effect of Pakistani ethnicity (versus White British ethnicity) after adjusting for a variety of environmental factors on the risk of EBV seropositivity in the first two years of life (OR 2.16) ^21^. This pattern is also seen for tonsillar EBV infection - among tonsils extracted from healthy individuals at various ages, the prevalence of EBV DNA increased monotonically with age for males (peak 77.4% for >35 year-olds), but reached a peak at 79.3% for females from 15-24, and remained at that plateau^22^.

Several large studies in other populations have demonstrated a decline in age-adjusted seroprevalence of EBV over the past 15 years ^4–6^. Interestingly, our study demonstrated consistently higher EBV seroprevalence among all age brackets compared to a comparable older study in similar cohorts^18^-this may reflect a genuine increase in seroprevalence in the UK, differences in population sampling, or different sensitivity and specificity of the assays used (our study used VCA whereas previous studies have used EBNA). In support of the first possibility, our estimates closely mirror those from a recent random sampling of UK Biobank participants: among 40-69 year olds in UK Biobank, EBV seroprevalence was 94.7%^1^. In our study, among the oldest age bracket EBV seroprevalence was 96.0%. A weakness of the HES dataset is that the incidence of all diseases appears to be increasing due to improved coding; we have been careful not to over-interpret this finding.

Understanding the temporal trends and host determinants of EBV serostatus are essential for rational vaccine design and deployment. In principle, vaccination against EBV could help to reduce rates of EBV-associated malignancies and EBV-associated autoimmune diseases. Although there is still no effective vaccine available, several are in development, and phase 2 results from a recombinant gp350 vaccine suggested an effect on IM rates without affecting overall seroprevalence ^12,23–25^. The population-level efficacy of such a vaccine depends crucially on the age at which it is administered, with earlier vaccines predicted to offer greater overall reductions in rates of IM and other EBV-related sequlae ^11^. Our empirical data showing high seroprevalence rates even in the youngest age bracket support the modelling data, suggesting that, in terms of maximising efficacy, EBV vaccines should be delivered in the first few years of life.

The risk of primary EBV infection manifesting as IM depends crucially on the age of the individual, with a higher risk at higher ages^26^. It follows that risk factors for IM include risk factors for delayed primary EBV infection, such as low household crowding, high levels of sanitation, low levels of intimate contact with others, and ethnicity^26,27^. In addition to environmental risk factors, host genetics influence an individual’s risk of IM - the largest GWAS to date identified a single locus in the MHC I region which had a modest effect (OR 1.08) on IM risk^28^ Understanding the risk factors for IM is challenging for several reasons: it is poorly coded in secondary care databases (as the majority of cases are not severe enough to lead to hospital admission), it is challenging to diagnose clinically as the presentation can be non-specific and most proposed risk factors confound one another (e.g. deprivation, crowding levels and hygiene levels are all clearly interrelated and depend on living environment). In our study, we use prevalent cases from a large East London cohort to examine the effects of ethnicity, BMI, smoking, and deprivation. We demonstrate that being white, from less deprived backgrounds, being overweight, and not smoking are associated with a higher risk of IM. Although the effects persist after correcting for confounding in a combined model, we cannot rule out that these exposures are in fact all proxies for lower levels of household crowding or other early life determinants. Intriguingly, we find that the effects of smoking and BMI differ between ethnicities. This may be explained by these traits being proxies for different exposures between different ethnicities (e.g. theoretically, being overweight could be a marker of high wealth in one group and deprivation in another). Alternatively, this may reflect a genuine biological effect whereby BMI and smoking interact with genetic background to influence IM risk. It is important to note that our study used prevalent cases, which prevents us from accurately determining the timing of exposures relative to the timing of IM, and introduces the problems of reverse causation, confounding, and recall bias.

A major concern regarding EBV vaccines is that these vaccines disrupt the host-pathogen interaction and have deleterious consequences for host and population immunity. EBV achieves long-term immune evasion by transforming naive B cells into a resting memory B cell phenotype and activating latency programmes. Given this tropism, and the widespread prevalence of EBV, there is an evolutionary argument that humans have co-evolved with EBV due to a selective advantage offered by the virus. Specifically, it is possible that EBV infection promotes herd and individual immunity to a broad range of pathogens by expanding and maintaining the size of the memory B cell pool. A theoretical concern that emerges in response to this argument is that vaccination against EBV have unintended consequences for B cell memory at an individual and population level. Our pilot data showing that EBV-seronegative individuals do not have lower levels of circulating antibody to common vaccine antigens and pathogens is reassuring, and will need to be replicated on a larger scale prior to allay this theoretical concern.

In summary, our data suggest that, in the UK, EBV seroconversion is taking place earlier in life, that overall EBV seroprevalence is increasing and remains very high (>95% of 20-25 year olds), IM incidence is increasing, and there are several environmental exposures associated with IM risk (ethnicity, deprivation, smoking, and BMI). Furthermore, we provide preliminary evidence that EBV seronegative persons do not have lower levels of circulating antibodies to common pathogens and vaccine antigens.

## Data Availability

Code on Github: https://github.com/benjacobs123456/benjacobs

## Acknowledgements

We are grateful to Barts Charity for supporting this work, to the SEU for providing sera, to Jaap Middeldorp for providing anonymised EBV-seronegative sera, to Mark Jitlal for his help in data extraction from the ELGP database, and to all the volunteers and patients who have provided their data for the study.

---

**Supplementary table 1:** demographics of EBV-seropositive and EBV-seronegative individuals included in the seroprevalence study.

|  | *Negative (n, %)* | *Positive (n, %)* |
| --- | --- | --- |
| ***Overall n*** | 343 (14.75%) | 1982 (85.25%) |
| ***Age band*** |  |  |
| *0 to 5* | 88 (30.03%) | 205 (69.97%) |
| *5 to 10* | 98 (23.9%) | 312 (76.1%) |
| *10 to 15* | 84 (19.13%) | 355 (80.87%) |
| *15 to 20* | 48 (8.62%) | 509 (91.38%) |
| *20 to 25* | 25 (3.99%) | 601 (96.01%) |
| ***Sex*** |  |  |
| *F* | 161 (13.47%) | 1034 (86.53%) |
| *M* | 182 (16.11%) | 948 (83.89%) |
| ***Location*** |  |  |
| *Exeter* | 33 (15.71%) | 177 (84.29%) |
| *Manchester* | 275 (15.61%) | 1487 (84.39%) |
| *Newcastle* | 34 (11%) | 275 (89%) |
| *Royal London* | 1 (2.27%) | 43 (97.73%) |

**Supplementary table 2:**
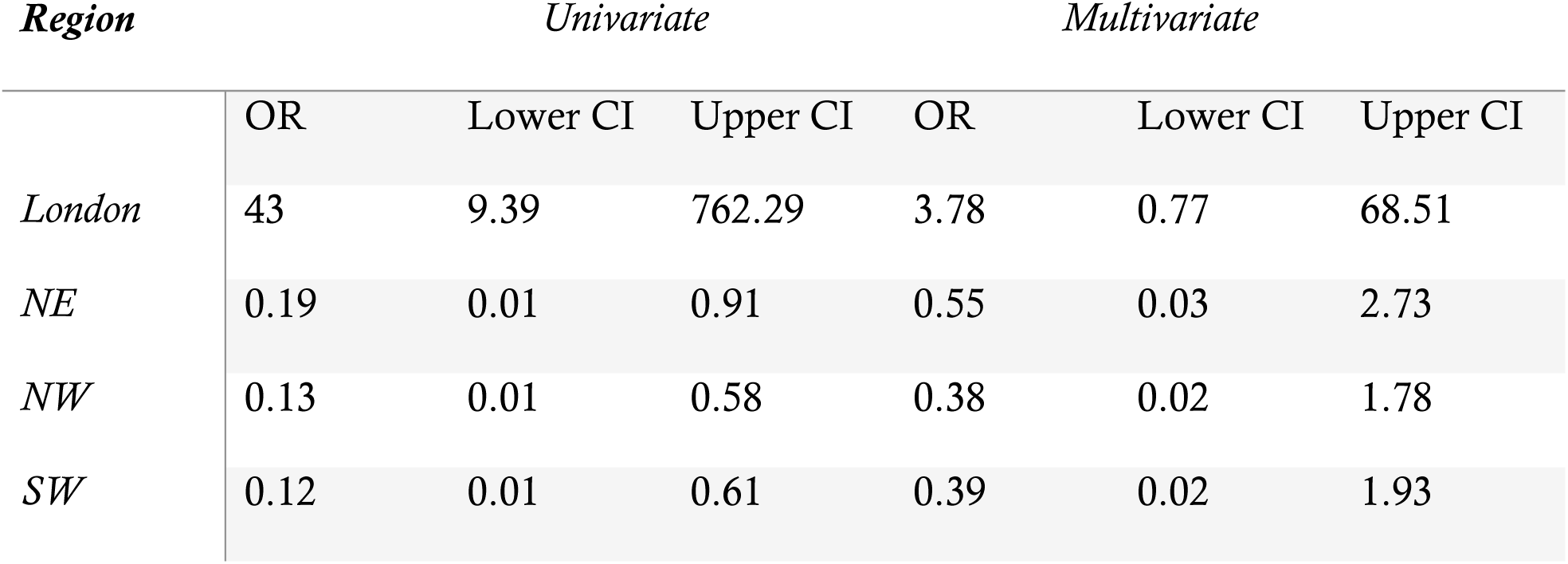
Effect estimates from univariate and multivariate (adjusted for age and sex) logistic regression modelling EBV seropositivity using region as the exposure.

**Supplementary table 3:** raw incidence of HES-coded IM (cases per 100,000 individuals) among males and females in three time windows.

|  | MALES |  |  | FEMALES |  |  | F:M RATIO |  |  |
| --- | --- | --- | --- | --- | --- | --- | --- | --- | --- |
|  | 2002-05 | 2006-09 | 2010-13 | 2002-05 | 2006-09 | 2010-13 | 2002-05 | 2006-09 | 2010-13 |
| **0-4** | 4.31 | 5.23 | 4.94 | 2.88 | 2.85 | 3.14 | 0.67 | 0.54 | 0.64 |
| **5-9** | 4.52 | 4.73 | 6.19 | 3.67 | 4.14 | 5.64 | 0.81 | 0.88 | 0.91 |
| **10-14** | 6.09 | 7.2 | 7.7 | 10.96 | 10.58 | 11.64 | 1.80 | 1.47 | 1.51 |
| **15-19** | 30.51 | 35.34 | 36.1 | 33.85 | 41.44 | 44.34 | 1.11 | 1.17 | 1.23 |
| **20-24** | 15.29 | 17.85 | 20.32 | 8.09 | 10.5 | 13.95 | 0.53 | 0.59 | 0.69 |
| **25-29** | 5.96 | 5.97 | 6.8 | 2.63 | 3.05 | 4.01 | 0.44 | 0.51 | 0.59 |
| **30-44** | 2.09 | 2.01 | 2.27 | 1 | 1.21 | 1.27 | 0.48 | 0.60 | 0.56 |
| **45-64** | 0.53 | 0.67 | 0.93 | 0.38 | 0.44 | 0.67 | 0.72 | 0.66 | 0.72 |
| **65-74** | 0.27 | 0.28 | 0.63 | 0.1 | 0.33 | 0.49 | 0.37 | 1.18 | 0.78 |
| **75+** | 0.13 | 0.39 | 0.32 | 0.12 | 0.15 | 0.17 | 0.92 | 0.38 | 0.53 |

**Supplementary table 4:** likelihood ratio p values comparing models of IM risk without pairwise interaction and with pairwise interaction between the named exposures. P values below the Bonferroni-adjusted threshold of 0.008 are highlighted in bold.

|  | *BMI* | *Smoking* | *IMD* | *Ethnicity* |
| --- | --- | --- | --- | --- |
| *BMI* |  |  |  |  |
| *Smoking* | 0.421 |  |  |  |
| *IMD* | **<0.005** | 0.112 |  |  |
| *Ethnicity* | **<0.0001** | **<0.0001** | **<0.0001** |  |

